# Barriers and enablers to accessing HIV services at South Sudan military facilities: Using qualitative data for program improvement

**DOI:** 10.1101/2023.11.15.23298584

**Authors:** Ally A. R. Lasu, Awin Changjowk, Shanice Fezeu Meyou, Habib D. Awongo, R. Craig Lefebvre, Justin Tongun, John Woja Elinana, Lauren Pindzola Courtney

## Abstract

**Introduction:** South Sudan established an HIV program for the South Sudan People’s Defense Force (SSPDF) in 2006, recognizing the potential national security threat posed by HIV’s impact in post-conflict settings. By 2018, the SSPDF program’s scope had expanded to include three VMMC clinics and four ART service delivery facilities. This qualitative study examined participant feedback on two existing HIV services, ART and VMMC, with the goal of identifying and prioritizing barriers and improving program performance. We used focus group discussions to gather information from male and female participants chosen at random across the four main project sites.

**Methods:** We conducted a cross-sectional qualitative study from March to April 2022 involving 177 people (108 men and 69 women) within fifteen focus group discussions. We collected data on enablers and barriers to HIV service uptake and utilization using a thematic framework approach.

**Results:** Perceived high-quality HIV services in a secure environment, organizational support systems, peer-led mobilization, and transportation facilitation were all identified as HIV service delivery enablers. HIV service delivery was hampered by knowledge gaps, poverty and food insecurity, access issues, a lack of treatment support groups, transportation challenges, social cultural barriers (stigma and discrimination), and the need for permission from commanders.

**Conclusions:** The findings show some overlap in client needs for VMMC and ART services. To increase ART retention and overall HIV service demand, the program will focus on improving the quality of HIV services and tailored client support, as well as addressing barriers resulting from structural and social cultural challenges to increase ART retention and overall HIV service demand.

## INTRODUCTION

In 2011, the Republic of South Sudan proudly became the world’s newest nation and Africa’s 55th country. Since then, development efforts in South Sudan have slowed because of recurring internal conflict, insecurity, and the COVID-19 pandemic (1), leading to poverty, food insecurity, deficits in education and health systems, internally displaced people, and other post-conflict effects (2–7). Ethnic conflicts in 2013 and 2016 have led to armed groups targeting civilians, destabilized new establishments, and displaced people (2–5, 8). The economic crisis and disruption in farming led to famine in many parts of the country. Additionally, reduced harvest, cattle theft, poaching, and flooding left more than 8 million people food insecure in 2019 (3, 4). Gender-based violence against women has been fueled by this ongoing instability, violence, and displacement. More than 4 million people remained displaced in 2022, which directly affects livelihood, education, social protection, and access to health services (6). Although recent developments aim to strengthen the health system, preventable/treatable communicable diseases such as HIV pose as a major threat and a leading cause of death in South Sudan. (7, 9–12). As South Sudan recovers from years of conflict, this fragile setting impacts the communities, the health system’s infrastructure, and their capacity for prevention, surveillance, and treatment, signifying an imperative leap toward peace (12).

The South Sudan National Strategic Plan for HIV/AIDS (2021–2023) (13) prioritized increased access to quality HIV care, treatment, treatment as prevention, and TB/HIV collaboration. South Sudan’s HIV epidemic is classified as low and generalized with a prevalence rate of 1.9 among adults aged 15-49 (14); however, pockets of HIV are concentrated among key and vulnerable populations, where the prevalence is 5% or higher (15). Although globally and regionally new HIV infections are decreasing and treatment is increasing (16), in South Sudan there are an unknown number of people living with HIV (PLHIV) who are virally suppressed and only an estimated 35% of people living with HIV know their status (14). The Ministry of Health (MoH) and related bodies have the national mandate of overseeing health care delivery in South Sudan. The unique HIV threats faced by the military, on the other hand, necessitate a contextualized forces policy to fill gaps left by the national program.

In 2006 South Sudan established an HIV program for the South Sudan People’s Defense Force (SSPDF), recognizing the potential national security threat from HIV’s impact. The HIV Secretariat oversees the program, which is tasked with planning, developing, and implementing quality HIV prevention, care, treatment, support, and impact mitigation services for military personnel, their families, and host communities. The military HIV program addresses institution-specific issues such as military personnel culture, access, deployment patterns, and the general work environment. The program has evolved from a narrow focus at Juba Military Hospital (JMH) in the capital city to a more expansive HIV care and treatment program, opening clinics in Wau Military Hospital (WMH) and Malual Chaat Military Hospital (MCH) in 2018 and 2021, respectively. Voluntary medical male circumcision (VMMC) services were introduced in JMH in 2018 and expanded to WMH and MCH in 2021. Despite these achievements, there is still a significant unmet need for HIV services among the military population (15).

Although ART and male circumcision are proven to be effective in reducing HIV-related morbidity, mortality, and transmission, addressing barriers impacting client access, program uptake, and coverage is critical to improve services, activities, stigma, and inequity (17). Used as part of combined prevention, the effectiveness of male circumcision in reducing sexual transmission of HIV from HIV-positive women to HIV-negative men has been shown in randomized control trials conducted in Kenya (18), South Africa (19), and Uganda (20) and consequently widely accepted since 2007 (21). Due to these achievements, the Joint United Nations Programme on HIV/AIDS (UNAIDS) investment framework (22) recognized male circumcision as one of the “basic program activities” with proven effectiveness in HIV prevention in generalized epidemics with a low prevalence of male circumcision. Additionally, investments in HIV response increase access to ART, with the goal of eliminating HIV as a global public health threat by 2030 (23). This strategy uses an array of biomedical, behavioral, and structural approaches (e.g., sexually transmitted disease control, voluntary counseling and testing, harm reduction, mother-to-child transmission prevention, blood safety, infection control in health care, structural interventions, and HIV-treatment programs and medical male circumcision) to achieve maximum impact.

Factors such as marginalization and stigmatization continue to pose as challenges and barriers especially among vulnerable populations. Access to HIV services may be hampered or mitigated by a variety of factors that occur at the individual, health provider, or health system level(24). Findings from studies conducted in Kenya (25), Uganda (26, 27), and Malawi (28) showed that the barriers and facilitators to care linkage are multifactorial. Major barriers include a lack of social networks, economic vulnerability, and fear of the consequences of stigma. Overcrowded clinics, unwelcoming staff, and unintegrated facilities resulted in linkage failure at the program level. Socioeconomic support from networks of health providers, family members, and friends was a key facilitator of linkage to HIV services.

This paper takes a novel approach to complement other efforts to understand the barriers and enablers to HIV service delivery in South Sudan’s post conflict military settings. The qualitative study examined participant feedback on two existing HIV services, ART and VMMC, to identify and prioritize barriers and to improve program performance. For example ART initiation and retention in treatment numbers at 12, 24, and 60 months, and client circumcised and adverse events reported for VMMC. Prior to this study, the South Sudan military HIV program had been implemented for military personnel, their families, and host communities without factoring in client perspectives about service delivery. RTI International/SSPDF, with support from the Department of Defense HIV/AIDS Prevention Program (DHAPP), aims to make HIV and VMMC services available and accessible to the wider military population at substantial HIV risk within the project catchment areas. However, there is limited information on beneficiary population perspectives about HIV services in those locations. Understanding the perspectives and preferences of HIV service beneficiaries, especially ART and eligible VMMC clients, is essential to designing programs that meet clients’ needs. Additionally, few studies have described how they have used data to improve program performance.

These are not new programs; ART services have been available in the South Sudan military since 2008, and VMMC services have been available since 2018. Because effective HIV prevention and treatment services are now available, RTI plans to use the study findings to expand tailored socio-behavioral change services by developing a comprehensive demand creation plan to address military and host community acceptance of, and generate demand for, these public health interventions to increase ART retention and overall HIV service demand. The implementation of the demand creation plan will be based on our experience designing and implementing socio-behavioral change programs to promote uptake of health products and services, including ART and VMMC services, but will also draw on best practice tools developed by PEPFAR and UNAIDS. Before implementing and adjusting activities to respond to changing public health programming conditions, providers will use the resulting demand creation plan to define and understand local military community needs. Importantly, the demand creation plan will be updated on a regular basis to ensure that its components remain relevant and useful.

## METHODS

### Setting

In March 1-April 30, 2022, we conducted 15 mixed-gender focus group discussions (FGDs) to identify barriers and enablers to accessing routine ART and VMMC services provided at three of four project sites where clients present voluntarily for free HIV services: JMH, WMH, and MCH.

##### Definition of Terms Used: Enablers, Barriers, and Treatment Supporters

- **Enablers**, as used in this study, are strategies, activities, and approaches aimed at improving accessibility, acceptability, uptake, equity, quality, effectiveness, and efficiency of HIV interventions. Enablers operate at all levels of society—at individual, community, institutional, societal, and national, regional, and global levels.
- Barriers, as used in this study, are impediments to HIV service uptake, including structural, sociocultural exclusion and marginalization, stigma, and inequity. If left unaddressed, these barriers will undermine the provision of HIV services, especially for priority populations such as the military.
- **A treatment supporter** is any person (including a family or PLHIV support group member) who is willing to help play a vital role in guiding and motivating HIV patients during their care and treatment; they are accepted by the patient and routinely engage with the health services (29).

### Participant recruitment

The FGD study gathered qualitative data from five FGD groups at each of the three program sites. Three VMMC-focused groups included circumcised males, non circumcised males, and female relatives of soldiers. Two ART-focused groups included ART-stable and ART-unstable clients.

For each focus group session, we invited seven to nine potential participants targeting specific ages and genders to ensure representation, as guided by site-specific program data. RTI screened potential participants; those who met the study criteria and agreed to participate were assigned to the appropriate FGD sessions. We confirmed ART adherence status through client records at the facilities; we assessed male circumcision status through self-reporting. We obtained verbal informed consent, which was documented in the participant log and basic demographic data from all participants. Approval for the study was obtained by RTI’s Institutional Review Board.

### Data collection

We conducted the FGDs in English; a trained moderator translated them into local languages (Dinka and Juba Arabic) as needed. The moderator used a semistructured questionnaire to facilitate the discussions with open-ended and probing questions. A notetaker recorded responses in real time, using audio-recordings to provide partially transcribed notes and translate all notes into English; audio-recordings allowed us to review for quality assurance. We provided each participant refreshments and travel cost reimbursement of about $20USD.

### Data collection instruments

Standardized interview questions were developed for each type of respondent. Sample interview questions are shown below.

##### Sample Interview Questions

- How would you describe our services?
- Based on your experience with our services, would you return for services?
- What are three important feature that you think we are missing?
- In your opinion, what important services currently provided at the ART clinic would you like to see continue?
- What are your biggest challenges in attending services at SSPDF facilities?
- Do you have a clear understanding of how to navigate or find different service points around this facility?
- Do you think clients would like to be connected to peer support network? This would be a network of people taking ARVs like themselves to support and encourage them to ensure their treatment is not interrupted.

### Data analysis

We used an inductive analytic approach to identify and describe the barriers and enablers of HIV service delivery in the DHAPP program. Two researchers reviewed the FDG notes and audio-recordings to identify key themes related to the topic. They grouped the themes and examined the narratives of FGD sessions per location. A senior researcher independently reviewed the transcripts to assess consistency and accuracy of themes identified. Areas of inconsistency were discussed with the moderator, noting definitions and context. The researchers discussed the findings and wrote summary notes and placed themes into analysis tables. Final results were written to reflect all themes identified.

### Using qualitative data for program improvement

Program leadership used the FGD data to identify program successes, needs for program improvements, and solutions. Additionally, leadership validated the outcomes with other available data sources such as routine program site visits, standard project listening sessions with SSPDF, and in-depth reviews with the implementation team. Integrating military perspectives into decision-making at all stages of programming planning and implementation is a key element of effectively using these data sources.

We examined FGD-identified barriers using a two-factor model to prioritize which barriers to address first. First, “*Is this barrier within our control to solve?*”; in other words, does the solution fit into the project scope, is there sufficient funding, is this reasonable to address within the period of the project? Second, “*To what extent does this impact clients receiving services?*” We used multiple data points to determine client impact, including the extent to which focus group participants discussed this issue, if multiple sites were impacted, and if this was an issue for both ART and VMMC.

## RESULTS

All 177 clients recruited at the three sites were eligible, and all consented to participate, as shown in Table 1.

**Table 1.** FGD Participants by Gender and Site.

| Facility Name | Males | Females |
| --- | --- | --- |
| Juba Military Hospital | 44 | 23 |
| Wau Military Hospital | 24 | 26 |
| Malual Chat Military Hospital | 40 | 20 |
| *Total* | *108* | *69* |

We describe the specific ART and VMMC enablers and barriers below, in addition to participant perspectives based on their interaction with the program.

### Enablers

#### Staffing (ART and VMMC)

Overwhelmingly, both ART-stable and Interruption in Treatment (IIT) participants from all three sites expressed gratitude for the welcoming nature of the ART providers. JMH clients in particular were more likely to comment on the welcoming nature of its staff; JMH has been in operation the longest. Participants particularly valued their providers’ openness, honesty, and level of confidentiality when offering HIV services. A current JMH ART client noted, “*Doctors have a good approach to clients and offer words that calm my heart. Seeing that I have no help from my children which is a challenge*.”

VMMC participants pointed out that the staff at the VMMC clinics treat all their clients with dignity and respect. They ensure that clients feel welcome and receive health education while addressing language difficulties as needed and supporting clients to utilize all the available services. When asked about their experience and whether they would return for additional services or recommend them to others, Bor (MCH) participants responded: “*Yes, I will come back because services are good and to bring other people because I know this place”, “Yes, environment is good, doctors are polite and quality circumcision service.”, and* “*Yes, I am likely to recommend these services for it is because of good quality, free of services, and free transport.”*

#### Peer/social support (ART)

ART participants frequently mentioned receiving emotional or material support from a support group, family member, friend, or a community health worker. Participants cited supporters’ traits such as trustworthiness, availability, and good communication as factors beneficial to a successful partnership. Some peer supporters helped participants by teaching them how to navigate the health system and encouraging them to keep track of their pill count and clinic appointment dates. A participant at WMH shared her experience with a community health worker: “*A lady took my phone number, and then checked on me – I am happy. Usually when I don’t show up, they call to ask me why I have not come for medication. Which is good*.” In these settings where facility staff numbers are inadequate, community health workers are critical in addressing patient challenges including psychosocial and adherence issues.

ART participants in MCH and WMH mentioned the need for treatment supporters. One participant noted that the “*Support group can help me when I am frustrated or missed an appointment.*” Another client highlighted the psychosocial support: “*The group will check and encourage each other to relieve thoughts and stress*.” Other clients also emphasized the importance of treatment supporters in treatment adherence *“…can help me through talking to me and in taking medicine*.” Several participants echoed the idea of peer supporters providing life skills, hope, and a positive outlook on life to clients. Such interventions increase clients’ individual and social motivation, resulting in the expansion of support networks and the promotion of optimism and positive living. Another participant reiterated the type of support they received from a health worker: “*There was a [health provider], he had a vehicle. He would call and if you are very sick, he would come looking to check on you and bring your medication.*” Such community outreach clearly contributes to enhanced HIV service delivery quality and uptake and by extension the dignity and quality of life of the clients.

#### Organizational support (ART)

The program provides mobile phone–based reminders of scheduled HIV appointments for clients. Other support available for clients includes multimonth dispensing, community tracing, and relinkage of clients who experience interruption in treatment. Most participants appreciated receiving reminder and follow-up calls for their appointment or picking up their antiretroviral (ARV) refill medication. Tracking and following up of IIT clients and counseling are critical approaches that clients agree are necessary for patients to continue with their treatment. A client added: “*They contact you by phone to ask you why you have not come. They also send someone to look for you at your residence in case you don’t show up*.”

#### Client transport facilitation (VMMC)

Participants shared that the provision of transport to and from the facility saved them money.

#### Safety (VMMC)

Participants expressed surprise that they felt safe within the clinic, even sitting together with other clients from the diverse ethnic and political backgrounds that had been in conflict. Because of the 2016 civil war in the MCH area, participants noted their fear of leaving the safety of their UN compounds to come to the clinic. One woman had never left the UN compound, noting that the facilitator convinced her to come to the FGD; she was amazed that she could walk freely through town. A participant commented that the VMMC clinic was “*peace building*” between two tribes through their accessing services together. Despite the prevailing ethnic tensions after the 2016 conflict, participants reported that they felt no discrimination or fear of being attacked or harmed during the whole process.

**Peer-led mobilization (VMMC)** as a component of the VMMC intervention including community mobilization was valued by clients. One participant in Wau stated: “*I mobilized more than 15 clients for circumcision to come to this clinic because the services are good, and I will continue to do so.*”

### Barriers

Barriers, as used in this study, are impediments to HIV service uptake, including structural, sociocultural exclusion and marginalization, stigma, and inequity. If left unaddressed, these barriers will undermine the provision of HIV services, especially for priority populations such as the military.

#### Funding for transport (ART)

Many participants agreed that addressing the transportation challenge would improve access and minimize client dropout. A client said: “*If there was a way for easing the transport challenge, patients would be helped to reach their drug pick up location. This is because if you don’t have money for transport, you will not come to pick your medication.*” This was particularly noted in Juba.

#### Food insecurity (ART)

Most participants believed that providing food to PLHIV would improve treatment numbers. They shared that lack of food is a barrier to consistently taking medication. One client stated: “*One day I took my drugs when there was no food and I felt dizzy, so I stopped taking the drugs*” and added “*I stop taking my drugs every time I do not have food.*” The practice of delaying ART medication because of lack of food was a recurrent theme in some groups. According to the participants, taking ARV drugs with juice was requested in multiple groups to help the pills go down more easily.

Female participants were more vocal about this problem. They highlighted the lack of employment opportunities and suggested being given business loans which would empower them to set up businesses to address the food insecurity problem in the long term.

#### Stigma and discrimination (ART)

Nearly all participants discussed perceived community stigma as a barrier with extreme community-level stigma noted at MCH; comments on this topic were lengthy. Participants described how stigma undermines one’s identity and capacity to cope with the disease. Multiple participants reported hiding their medications for fear of household members discovering they were HIV positive. Facilitators commented that the WMH and MCH sessions for IIT participants were emotionally touching; all participants’ stories were difficult to hear. Women spoke up more than men and told their truth.

One participant had delayed HIV testing because of fear, and thus, treatment was delayed. Fear of discrimination and of potential rejection within his immediate family and community prevented his disclosure to even his close family. He said: “*This disease is top secret, and no one must know at all apart from doctors.*” In MCH, a participant reported that he may not want to keep or take his medications at home for fear of being discovered by family. “*I fear taking medication when other people are present in the house.*” Learning that one is on ART medication may create hostility from family members who hold negative attitudes and beliefs about HIV, potentially leading to disruption in family harmony.

#### Stigma/social cultural attitudes (VMMC)

All VMMC participants agreed that they knew about VMMC, but some specifically emphasized the sociocultural barriers to adopting the practice.

One participant noted: “*I know well about medical circumcision but…Dinka Bor culture does not allow male circumcision, despite it being practiced in town.*” Another participant stated that: “*I’m married and for me to get circumcised, my wife must agree before I do it and will not be circumcised if my wife refuses.*”

One participant reflected the reason for his hesitation: “*Male circumcision should be agreed to by our female counterparts to avoid divorce. A wife of my friend divorced after his circumcision.*” Another expressed fear and stigma: “*I’m aware about the service and its importance, but I’m not ready to be circumcised, fearing community abuse. In my community circumcision is not allowed and if one does it, you and your children will be abused until your death.*”

#### Knowledge gaps (ART and VMMC)

Confusing VMMC with traditional circumcision, some participants only know the traditional circumcision that is conducted by traditional healers within the villages under poor hygienic conditions. So, they confuse VMMC with that approach, leading to some men and women expressing unfounded fear of incompetent providers and potential adverse events such as bleeding, swelling (keloids), and pain after medical circumcision. Participants on ART emphasized the lack of routine counseling to address knowledge gaps about the ART’s side effects. Some participants even inquired about the availability of injectable ART and single-dose medication for children in South Sudan.

#### Limited access (ART and VMMC)

Some participants highlighted lack of access to VMMC services within their localities. Available services are few and are located within military garrisons, which for some may be far from where military and host communities reside. This usually impacts access because of high costs or limited transport options.

For ART services, IIT participants indicate the long distance to the clinics, and other access issues, prevented them from seeking regular treatment. This was true for participants newly on treatment who cannot benefit from community refills.

### Using Data for Program Improvement

We summarize enablers and barriers in Tables 2 and 3, indicating which focus group clients (VMMC or ART) remarked on the issue and (for Table 3) if resolution of this issue was within the project’s control.

**Table 2.** Summary of Enablers Key for ART and VMMC Clients in South Sudan.

| Order of Priority | Enablers | Description | ART | VMMC |
| --- | --- | --- | --- | --- |
| 1 | Staffing | Positive experiences with welcoming clinic staff | ✓ | ✓ |
| 2 | Peer/social support | Peer support groups available | ✓ |  |
| 3 | Organizational support | Client support such as appointment reminders, multimonth dispensing, community-tracing and relinking | ✓ |  |
| 4 | Client transport facilitation | Transportation to the clinic is provided for clients |  | ✓ |
| 5 | Safety | Clients feel safe at the clinic |  | ✓ |
| 6 | Peer-led mobilization | Clients share positive experiences with peers, encouraging them to join |  | ✓ |

**Table 3.** Model for Prioritizing Barriers for ART and VMMC Clients.

| Priority Groups | Barrier | Problem | ART | VMMC | Within Project Control |
| --- | --- | --- | --- | --- | --- |
| 1 | Knowledge gaps | Myths and misconceptions | ✓ | ✓ | ✓ |
|  | Access issues | Limited service points | ✓ | ✓ | ✓ |
| 2 | Support groups | Lack of strong, organized PLHIV groups that can contribute to reduction of stigma within their communities | ✓ |  | ✓ |
|  | Treatment support | Clients across ART sites still struggle with access to treatment supporters | ✓ |  | ✓ |
|  | Transport | Transport costs | ✓ |  | ✓ |
| 3 | Permission from commander | Commanders must provide permission since soldiers must miss work |  | ✓ | ✓ |
| 4 | Sociocultural | Stigma | ✓ | ✓ | ✗ |
|  | Safety | Clients' fear of leaving the safety of their UN compounds because of the 2016 civil war |  | ✓ | ✗ |
|  | Poverty and food insecurity | Women empowerment to address family nutrition | ✓ |  | ✗ |

## DISCUSSION

As a nascent post-conflict country, South Sudan’s myriad infrastructure, human resource, and structural challenges plague public service delivery, especially health services. Findings from our focus groups identified enablers and barriers to HIV service delivery, allowing our team to prioritize strategies to improve program performance.

Enablers of HIV service delivery include the perceived high quality of services, satisfaction with the providers and services, and availability of transport. This feedback shows that participants hold positive relationships with their providers; the human touch is critical for driving client demand for services and creating a friendly facility. Because some of these clients lack peer support, they are less likely to engage in HIV care, and thus program facility staff become a valuable resource for client engagement and adherence to care. Staff at the facility can assist clients in making sense of confusing health information, provide psychosocial support, influence their treatment-related behaviors, navigate the health care system, and improve relationships with peers in their communities.

Maintaining free-of-cost HIV services, clean workspace, staff cooperation, privacy and confidentiality, and facilitating client transport, safety, and security, were stated as perceived indicators of the quality of HIV services offered at the three facilities. Thus, improving demand for HIV services may require addressing these factors. A welcoming environment, good communication, and respect for patients should be implemented in all the project facilities to promote these outlined socially acceptable practices. There is also need for integration of services at all sites to provide opportunities to deliver interventions across the spectrum of needs of HIV-infected military men and women and the affected communities. We also noted the willingness with which the potential clients from different ethnic groups/tribes agreed to be booked, meet at scheduled central meeting points, and be transported together at zero cost to the VMMC site as a novel approach to promote peace. There is no reported case of discrimination against any client based on their tribe, ethnicity, religion, or political affiliation as they navigate the program health facilities.

We prioritized barriers by grouping them into four categories according to results shown in Table 3. Facilitator feedback noted the emotional strain participants expressed and difficulty they felt while conducting these FDGs; these topics are all critical for this extremely needy and vulnerable population. Although the team wished to address all issues highlighted, we prioritized them to enable positive changes moving forward.

**1- Top priority** factors that affect both ART and VMMC services and are within control of the program include **knowledge gaps** and **service access issues**.

### Knowledge gaps

HIV/AIDS knowledge is low in South Sudan because of low literacy rates (34.5%) among both men and women, particularly among those in the military (15). Traditional print and broadcast media that could be used to increase HIV awareness, such as radio, television, and new online media, are not accessible to a large portion of the population, both rural and urban. Furthermore, at the national and community levels, South Sudan lacks strong and diverse HIV champions to drive the HIV awareness and a mobilization agenda. Therefore, the program will ensure that clients have social connections or “buddies” who are involved in providing accurate and reliable HIV treatment information to clients in their health journeys and to hold them accountable.

**Access barriers** to ART services have been reduced in key regional settings by establishing facilities in military-rich areas, with a plan to further decentralize services to address unequal access to ART in the surrounding areas. Some respondents reported that affordability barriers are significantly higher for clients in more remote settings. This finding is consistent with South Sudan’s current HIV service delivery policy emphasis on strengthening community ART service delivery.

**2- High priority** factors affect only ART services; these include **support groups**.

### Support groups

Although the program allows stable PLHIV to meet once a month to share treatment experiences and support one another, it lacks self-formed support groups in larger communities to follow up on and address their unique individual ART challenges.

Although the military program has more than doubled its ART enrollment from 1,172 in 2019 to 2,597 in 2022, low treatment retention rates, which are currently less than 50% for the program, threaten to undo the program’s gains in ART enrollment over the last 8 years. Attrition is often seen as the result of inadequate patient preparation, which causes treatment interruptions, and a lack of strong community-level support for new PLHIV. In 2020, the MOH and its partners developed guidelines and protocols while keeping structural constraints of patients and the health system in mind. This protocol gives the client the ability to collaborate with health providers, institutions, and others to improve their own care. Improved adherence, increased motivation and confidence, and mutual support, as observed by Sabin (30), result in higher levels of physical, psychological, and social well-being and improved health outcomes (30).

**3- Medium priority** factors affect VMMC only; these include **transport** and **permission from commanders**.

### Transport

Similar to Ancia (31) findings, some participants were grateful for the free transport provided to the clinic for VMMC because otherwise they would need to walk long distances from their homes to the service point, since transportation is unavailable or too expensive (31). ART participants do not have access to transport, although the program provides community refills for stable clients.

### Permission from commanders

The military is a hierarchical institution in which subordinates need permission from their commanders to seek treatment on or off the military base for extended periods of time. For a service like VMMC, where clients need about 3-4 weeks to fully recover, military leaders must be convinced that VMMC is critical for their troops. Potential clients have been denied leave to attend such services in some cases where commanders do not understand their roles and have not been sensitized about their troops’ rights, including the right to life and health, and other human rights. The program continues to hold regular commander training sessions to ensure that new commanders are familiar with the HIV program, their roles in HIV service uptake and utilization, and that they remain accountable for the overall health of their troops.

**4- Low priority** factors are beyond the remit of the program; these include **safety, poverty/food insecurity, and stigma/sociocultural issues**.

### Safety

Since the conflicts in 2013 and 2016, approximately 1.96 million South Sudanese have been internally displaced, with the majority residing in UN-managed civilian sites. Food insecurity continues to affect large segments of the South Sudanese population because of the economic downturn, civil insecurity, the lingering effects of floods, and the prolonged conflict.

**Poverty** is widespread, and most people rely on subsistence agriculture and humanitarian aid. The additional burden of managing a chronic illness in a resource-scarce environment makes PLHIV even more vulnerable and in need of support.

### Stigma

Myths and misconception are powerful drivers of stigma (16) toward PLHIV in South Sudan. This leads to discrimination expressed by loss of jobs, intimate partner violence, expulsion from homes, and denial of basic health and other services such as leasing house. RTI has been able to provide HIV services in the four project facilities that are not perceived as "stigmatizing" by clients and may serve as a "beacon of hope" for less stigmatizing attitudes and behaviors in the communities they serve. This was accomplished by collaborating closely with PLHIV clients around project facilities to help create an environment that provides patients with HIV with timely, appropriate, and humane HIV care services.

Sociocultural issues, especially gender roles and relationships, play a significant role in the transmission of HIV and perspectives on its prevention and treatment. Participants discussed the impact of HIV on their lives, their role in HIV prevention, and obstacles to accessing HIV services. A comprehensive, multidisciplinary, systemic approach that includes community-wide education and empowerment and modifying legal and social structures that contribute to the spread of HIV is suggested as a necessary addition to the current national response. Only by forging strong partnerships between the different actors in health, education, women and youth groups, and political, religious, and opinion leaders will the impact of HIV be reduced in South Sudan.

We will use the lessons learned to design interventions that are appropriate and acceptable to the beneficiary population; delivered by providers with relevant personal and community experience; and tailored to address sociocultural influences such as stigma and discrimination. The military leadership and RTI are committed to data analysis and use for real-time program course-correction, while engaging meaningfully with the beneficiaries. By continuously triangulating available information, we will improve service delivery and program content to reflect local realities. Maintaining close partnership with and education of military leadership is central to ownership and sustainability of the program.

## CONCLUSIONS/NEXT STEPS

This study sought to ensure that HIV ART and VMMC programming in South Sudan is based on evidence; the program will adapt to local circumstances and target military community needs while promoting program effectiveness. We created a tiered system to prioritize client needs based on the barriers/enablers we identified from two HIV services (VMMC and ART) and those issues within project control. Our findings highlight some overlap in the needs of clients VMMC and ART services, enabling us to target both services to increase demand for HIV services. South Sudan’s fragile post-conflict context is noteworthy and influences the needs of the HIV service delivery clients.

As next steps, we plan to use the FGD feedback to prioritize demand-creation interventions to address the factors above to increase ART and VMMC service uptake with several next steps:

- Creating a comprehensive HIV awareness model that addresses knowledge gaps and emphasizes service delivery point availability in the short term.
- Collaborating with the military program team to identify and nurture PLHIV support groups in each of the three ART program locations.
- In the long term, exploring ways to sustainably increase access to ART and VMMC service locations. The program will also reach out to military leadership to identify command channels for troops who want to undergo VMMC to obtain permission promptly.
- Continuing to reach out to relevant government officials, partners, and stakeholders who may have solutions to the issues of safety, poverty, and sociocultural issues.

## LIMITATIONS

This study has several limitations that should be considered in future program improvement studies. Since the participants in this study were not chosen at random, the findings may not be applicable to all beneficiaries/patients served by the facilities. Recordings of the focus groups were not transcribed or translated because of programmatic costs. Translations were carried out during the sessions, with notes taken in English. Some participants in the ART-unstable focus groups revealed serious health conditions, with some requiring hospitalization during or immediately following the sessions. This could have limited their ability to participate effectively in the discussions. Because of the nature of group dynamics, “group think” may occur in focus group settings, resulting in themes that would not have emerged as frequently if interviews were conducted individually. Some discussion points may have been “lost in translation” because of language barriers in some study locations, because sessions required real-time translation, response, and note-taking. Despite these limitations, the study findings suggest several practical and programmatic implications that can help the SSPDF program improve service delivery.

## Data Availability

N/A relevant data submitted with the manuscript.

## ACKNOWLEDGEMENTS

We acknowledge the support of the DoD HIV/AIDS Prevention Program (DHAPP), as well as the SSPDF HIV secretariat and technical review and feedback from Katie Grimes.

